# Development of a parent-reported screening tool for avoidant/restrictive food intake disorder (ARFID): Initial validation and prevalence in 4-7-year-old Japanese children

**DOI:** 10.1101/2021.05.11.21254533

**Authors:** Lisa Dinkler, Kahoko Yasumitsu-Lovell, Masamitsu Eitoku, Mikiya Fujieda, Narufumi Suganuma, Yuhei Hatakenaka, Nouchine Hadjikhani, Rachel Bryant-Waugh, Maria Råstam, Christopher Gillberg

**Affiliations:** Gillberg Neuropsychiatry Centre, Institute of Neuroscience and Physiology, University of Gothenburg, Gothenburg, Sweden; Department of Environmental Medicine, Kochi Medical School, Kochi University, Kohasu, Oko-Cho, Nankoku, Kochi, Japan; Department of Pediatrics, Kochi Medical School, Kochi University, Kohasu, Oko-Cho, Nankoku, Kochi, Japan; Faculty of Humanities and Sociologies, University of the Ryukyus, Nishihara, Okinawa, Japan; Athinoula A. Martinos Center for Biomedical Imaging, Massachusetts General Hospital, Harvard Medical School, Charlestown, Massachusetts, US; Maudsley Centre for Child and Adolescent Eating Disorders, South London and Maudsley NHS Foundation Trust, London, UK; Department of Clinical Sciences Lund, Lund University, Lund, Sweden

**Keywords:** Avoidant/Restrictive Food Intake Disorder, prevalence, screening, impairment, Japan Environment and Children’s Study

## Abstract

The prevalence of avoidant/restrictive food intake disorder (ARFID) in the general child population is still largely unknown and validated screening instruments are lacking. The aims of this study were to investigate the prevalence of children screening positive for ARFID in a Japanese birth cohort using a newly developed parent-reported screening tool, to estimate the prevalence of children with ARFID experiencing physical versus psychosocial consequences of their eating pattern, and to provide preliminary evidence for the validity of the new screening tool. Data were collected from 3,728 4-7-year-old children born in Kochi prefecture (response rate was 56.5%), Japan, between 2011 and 2014; a sub-sample of the Japan Environment and Children’s Study (JECS). Parents completed a questionnaire including the ARFID screener and several other measures to assess convergent validity. The point prevalence of children screening positive for ARFID was 1.3%; half of them met criteria for ARFID based on psychosocial impairment alone, while the other half met diagnostic criteria relating to physical impairment (and additional psychosocial impairment in many cases). Sensory sensitivity to food characteristics (63%) and/or lack of interest in eating (51%) were the most prevalent drivers of food avoidance. Children screening positive for ARFID were lighter in weight and shorter in height, they showed more problem behaviors related to mealtimes and nutritional intake, and they were more often selective eaters and more responsive to satiety, providing preliminary support for the validity of the new screening tool. This is the largest screening study to date of ARFID in children up to 7 years. Future studies should examine the diagnostic validity of the new ARFID screener using clinically ascertained cases. Further research on ARFID prevalence in the general population is needed.

## 1. Introduction

Avoidant/restrictive food intake disorder (ARFID) is characterized by an avoidance or restriction of the *range* of foods and/or the overall *amount* eaten that cannot be explained by weight and shape concerns seen in other eating disorders such as anorexia nervosa. A diagnosis of ARFID requires a clinically significant negative impact on weight, nutrition and/or psychosocial functioning. Lack of available food, culturally sanctioned practices, and other medical or psychiatric conditions should not adequately account for the eating disturbance. ARFID was added to the DSM-5 in 2013 as a feeding and eating disorder diagnosis (American Psychiatric Association, 2013) and is also included in the ICD-11 (World Health Organization, 2018). Despite a burgeoning body of research, the prevalence of ARFID in the general population is still largely unknown. Having relatively precise estimates of ARFID prevalence in the population is important to assess the impact of ARFID on the population and to appropriately organize health care. However, large epidemiological studies require a lot of time and effort, and most importantly, screening tools for ARFID need further development and validation (Eddy et al., 2019).

As of today and to the best of our knowledge, only eight studies of ARFID prevalence in the general population are available. Seven of these assessed self-reported ARFID symptoms; of which four studies included adults and three studies included children between 7 and 14 years of age. These seven studies yielded point prevalence estimates between 0.3 and 5.5% (Chen, Chen, Lin, Shen, & Gau, 2019; Chua, Fitzsimmons-Craft, Austin, Wilfley, & Taylor, 2021; Fitzsimmons-Craft et al., 2019; Hay et al., 2017; Hilbert, Zenger, Eichler, & Brahler, 2021; Kurz, van Dyck, Dremmel, Munsch, & Hilbert, 2015; Schmidt, Vogel, Hiemisch, Kiess, & Hilbert, 2018) (for details see Table 3 in (Dinkler & Bryant-Waugh, 2021)). The applied screening instruments differ significantly in number and type of questions and in their focus on assessing the diagnostic criteria (i.e., consequences of avoidant/restrictive eating and exclusion criteria) versus the drivers of avoidant/restrictive eating (e.g., lack of appetite, sensory sensitivity to food characteristics), which might explain the relatively broad range of prevalence estimates. Importantly, none of the screening tools has yet been validated against clinically ascertained diagnoses.

In younger children, self-reports of ARFID symptoms are not feasible, and *parent-reported* instruments are therefore needed. Only one study used a parent-reported questionnaire of ARFID symptoms to examine ARFID prevalence in younger children. The questionnaire consisted of five items covering main ARFID symptoms answered with *yes* or *no*, of which four items were used to identify ARFID (Gonçalves et al., 2019). ARFID symptoms were present in 15.5% of 330 Portuguese children aged between 5 and 10 years. Considering all other reported prevalence estimates, this estimate seems disproportionately high. The authors argue that the response format of the questions in combination with generally high concern of Portuguese parents about their children’s eating and weight might have led to an overestimation of the prevalence. Furthermore, no questions regarding the DSM-5 exclusion criteria were included in the parental questionnaire. In summary, the prevalence of ARFID in very young children in the general population is still completely unknown and there is a clear need for parent-reported screening tools in young children.

Another issue affecting ARFID prevalence rates is whether *physical* consequences of avoidant/restrictive eating (i.e., negative impact on weight, growth, or nutrition) are required to be present for diagnosis (as opposed to a negative impact on psychosocial functioning only). This is relevant as the way diagnostic criterion A is worded in the DSM-5 is somewhat ambiguous, which has led to some discussion as to whether DSM-5 criterion A4 (marked interference with psychosocial functioning) would be sufficient to meet criterion A in the absence of criteria A1-A3 which are related to the physical impact of avoidant and/or restricted eating (e.g., A1 - weight loss, A2 - nutritional deficiency, A3 - dependence on enteral feeding). In the upcoming DSM-5-TR, it will be clarified that *either* physical *or* psychosocial impairment (or both) is required for diagnosis, that is, criterion A4 alone is sufficient to meet criterion A (R Bryant-Waugh, personal communication, 1 April 2021), which is also the case in ICD-11 (World Health Organization, 2018). Criterion A4 might also be the criterion that is most challenging to assess, and its operationalization is not entirely clear (Eddy et al., 2019). Especially, in young children, this criterion might be difficult to evaluate, as parents and other caregivers often make wide-ranging accommodations to the child’s needs and wishes around food, so that the psychosocial functioning of the child might not be impacted significantly, while the psychosocial functioning of the *family* might well be. When assessing the criterion, it is therefore important to differentiate between consequences for the child versus for the family/caregivers. Concerns have also been raised regarding over-diagnosis of ARFID due to potential over-reporting of impairment by some parents on behalf of their children (Eddy et al., 2019). Previous studies on ARFID prevalence have not differentiated between physical versus psychosocial impairment through avoidant/restrictive eating and their impact on ARFID prevalence is not known.

Considering the lack of parent-reported screening tools for ARFID and the lack knowledge about ARFID prevalence in very young children and how it is affected by the interpretation of the DSM-5 diagnostic criterion A, the aims of this study were: (1) to investigate the prevalence of children screening positive for ARFID in a large birth cohort of Japanese children aged 4-7 years using a newly developed parent-reported screening instrument, (2) to examine the difference in ARFID prevalence depending on whether physical consequences of avoidant/restrictive eating are required or not, and (3) to provide preliminary evidence for the validity of the new screening tool. Considering previously reported prevalence estimates, we hypothesized that the ARFID prevalence in this sample would be less than 5%. We further hypothesized that the screening tool would differentiate children screening positive versus negative for ARFID on dimensions such as height, body mass index (BMI), restrictive eating, and overeating. Specifically, we expected children with ARFID to be shorter in height, to have lower BMI percentiles and to show more restrictive eating but less overeating.

## 2. Method

### 2.1 Participants

This study included a sub-sample of the Japan Environment and Children’s Study (JECS), an ongoing nationwide birth cohort study following approximately 100,000 children from pregnancy/birth until the age of 13. JECS includes 15 Regional Centers that recruited pregnant women via the collaborating local health care providers and local government offices where women registered their pregnancy. The Regional Centers were requested to cover more than 50% of pregnancies in the defined area of study (Kawamoto et al., 2014; Michikawa et al., 2018). Eligibility criteria for participation in the JECS are described in detail in Kawamoto et al. (2014). In collaboration with the Kochi Regional Centre of the JECS at Kochi Medical School we collected additional data in the Kochi cohort, a sub-cohort of the JECS including 6,633^1^ children born in Kochi prefecture between July 2011 and December 2014. A questionnaire was sent out to all parents in the Kochi cohort in December 2018. Responses were collected until 31^st^ October 2019. The response rate was 56.5% (n=3,746), an attrition analysis can be found in Supplement 1. This study was approved by the ethics committee at Kochi Medical School (ERB-102925 and ERB-104083). Participants gave informed consent before taking part in the study.

### 2.2 Measures

#### 2.2.1 Development of the ARFID screener

The questionnaire developed for this study is intended to screen for ARFID in children by parent-report. It was designed by three senior experts in feeding and eating disorders (RBW, CG, MR) with extensive experience in questionnaire development, and by a PhD student (LD). The items map closely onto the diagnostic criteria for DSM-5 ARFID and also examine the presence of the three drivers of food avoidance exemplified in the DSM-5: se*nsory sensitivity to characteristics of food, lack of interest in eating*, and *fear of aversive consequences of eating* (Thomas et al., 2017). Most criteria and the drivers of food avoidance were assessed with one item each, while two criteria were assessed with two items each. Table 1 shows items, response options, and required responses to meet the respective criterion. Criterion B (the eating disturbance is not due to lack of available food or a culturally sanctioned practice) was not assessed because we considered our cohort (a) affluent enough for food shortage to be relatively unlikely, and (b) culturally homogenous enough with no particular food restriction practice.

**Table 1.**
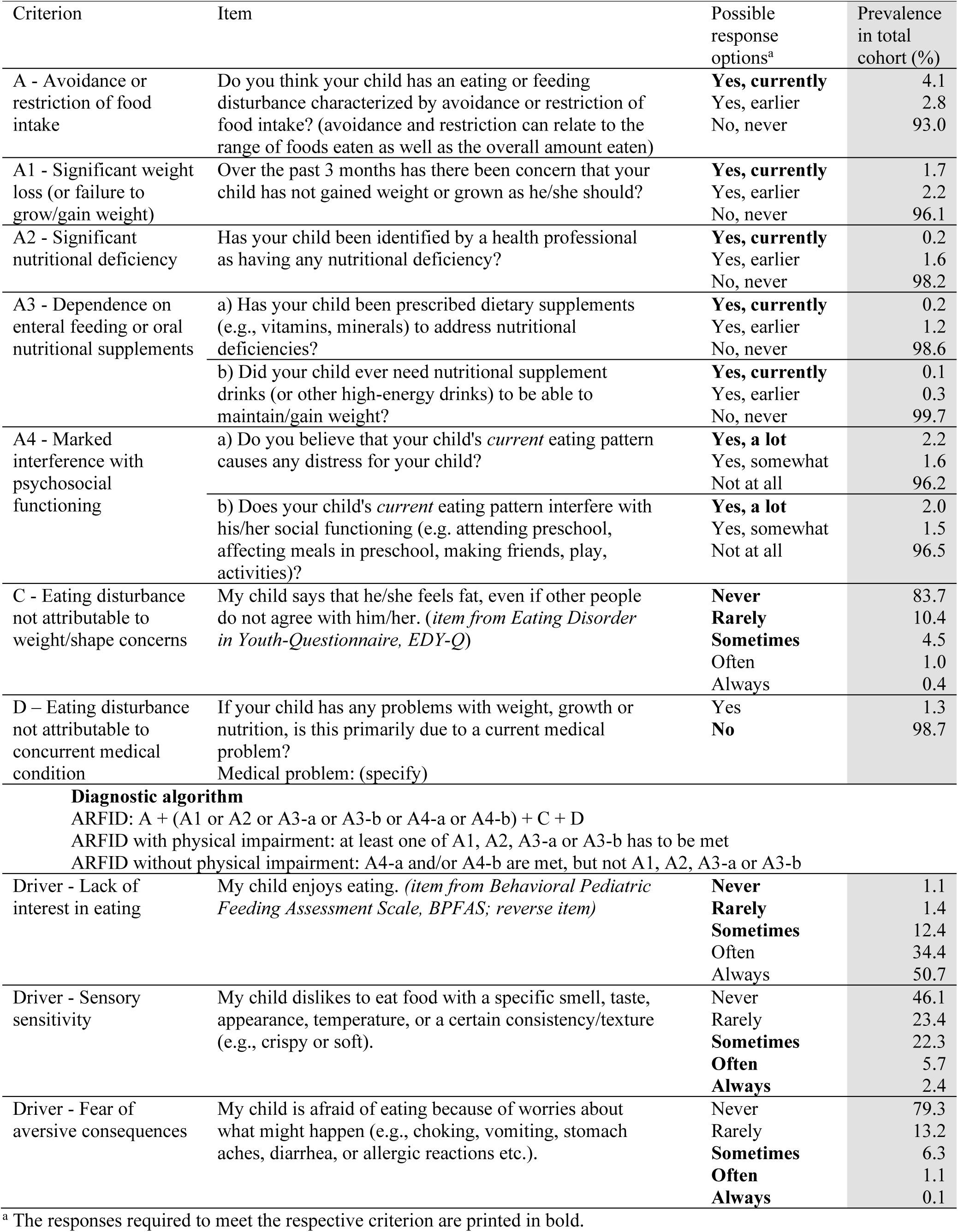
Items used to screen the DSM-5 diagnostic criteria for Avoidant/Restrictive Food Intake Disorder (ARFID), including response options, required responses for meeting the criteria, and prevalence of each response option in the total cohort

| Criterion | Item | Possible response options<sup>a</sup> | Prevalence in total cohort (%) |
| --- | --- | --- | --- |
| A - Avoidance or restriction of food intake | Do you think your child has an eating or feeding disturbance characterized by avoidance or restriction of food intake? (avoidance and restriction can relate to the range of foods eaten as well as the overall amount eaten) | **Yes, currently**<br>Yes, earlier<br>No, never | 4.1<br>2.8<br>93.0 |
| A1 - Significant weight loss (or failure to grow/gain weight) | Over the past 3 months has there been concern that your child has not gained weight or grown as he/she should? | **Yes, currently**<br>Yes, earlier<br>No, never | 1.7<br>2.2<br>96.1 |
| A2 - Significant nutritional deficiency | Has your child been identified by a health professional as having any nutritional deficiency? | **Yes, currently**<br>Yes, earlier<br>No, never | 0.2<br>1.6<br>98.2 |
| A3 - Dependence on enteral feeding or oral nutritional supplements | a) Has your child been prescribed dietary supplements (e.g., vitamins, minerals) to address nutritional deficiencies? | **Yes, currently**<br>Yes, earlier<br>No, never | 0.2<br>1.2<br>98.6 |
|  | b) Did your child ever need nutritional supplement drinks (or other high-energy drinks) to be able to maintain/gain weight? | **Yes, currently**<br>Yes, earlier<br>No, never | 0.1<br>0.3<br>99.7 |
| A4 - Marked interference with psychosocial functioning | a) Do you believe that your child's *current* eating pattern causes any distress for your child? | **Yes, a lot**<br>Yes, somewhat<br>Not at all | 2.2<br>1.6<br>96.2 |
|  | b) Does your child's *current* eating pattern interfere with his/her social functioning (e.g. attending preschool, affecting meals in preschool, making friends, play, activities)? | **Yes, a lot**<br>Yes, somewhat<br>Not at all | 2.0<br>1.5<br>96.5 |
| C - Eating disturbance not attributable to weight/shape concerns | My child says that he/she feels fat, even if other people do not agree with him/her. (*item from Eating Disorder in Youth-Questionnaire, EDY-Q*) | **Never**<br>**Rarely**<br>**Sometimes**<br>Often<br>Always | 83.7<br>10.4<br>4.5<br>1.0<br>0.4 |
| D – Eating disturbance not attributable to concurrent medical condition | If your child has any problems with weight, growth or nutrition, is this primarily due to a current medical problem?<br>Medical problem: (specify) | Yes<br>**No** | 1.3<br>98.7 |
| **Diagnostic algorithm** |  |  |  |
| ARFID: A + (A1 or A2 or A3-a or A3-b or A4-a or A4-b) + C + D |  |  |  |
| ARFID with physical impairment: at least one of A1, A2, A3-a or A3-b has to be met |  |  |  |
| ARFID without physical impairment: A4-a and/or A4-b are met, but not A1, A2, A3-a or A3-b |  |  |  |
| Driver - Lack of interest in eating | My child enjoys eating. (*item from Behavioral Pediatric Feeding Assessment Scale, BPFAS; reverse item*) | **Never** | 1.1 |
|  |  | **Rarely** | 1.4 |
|  |  | **Sometimes** | 12.4 |
|  |  | Often | 34.4 |
|  |  | Always | 50.7 |
| Driver - Sensory sensitivity | My child dislikes to eat food with a specific smell, taste, appearance, temperature, or a certain consistency/texture (e.g., crispy or soft). | Never | 46.1 |
|  |  | Rarely | 23.4 |
|  |  | **Sometimes** | 22.3 |
|  |  | **Often** | 5.7 |
|  |  | **Always** | 2.4 |
| Driver - Fear of aversive consequences | My child is afraid of eating because of worries about what might happen (e.g., choking, vomiting, stomach aches, diarrhea, or allergic reactions etc.). | Never | 79.3 |
|  |  | Rarely | 13.2 |
|  |  | **Sometimes** | 6.3 |
|  |  | **Often** | 1.1 |
|  |  | **Always** | 0.1 |

<sup>a</sup> The responses required to meet the respective criterion are printed in bold.

Children were identified with ARFID if the following criteria were met: (1) parents indicated that their child *currently* had an eating disturbance characterized by avoidance or restriction of food intake (criterion A), (2) the eating disturbance *currently* caused physical or psychosocial impairment for the child (criteria A1-A4, at least one of them had to be met), (3) the eating disturbance was not attributable to weight/shape concerns (criterion C), and (4) the eating disturbance was not attributable to a concurrent medical condition (criterion D). In addition, we differentiated between ARFID *with* physical impairment (i.e., negative impact on weight, growth, or nutrition; at least one of criteria A1-A3 had to be met) and ARFID *without* physical impairment (i.e., no negative impact on weight, growth, or nutrition; criterion A4 was met, but not criteria A1-A3).

As criterion A4 requires “*marked* inference with psychosocial functioning” (American Psychiatric Association, 2013), this criterion was considered to be met if at least one of the two items assessing this criterion was rated “Yes, a lot” (Table 1). For criterion C, we considered it sufficient to check for weight and shape concerns in order to exclude the possibility of the eating disturbance occurring “exclusively during the course of anorexia nervosa or bulimia nervosa” (American Psychiatric Association, 2013), since anorexia nervosa and bulimia nervosa are very unlikely to occur in this age group of 4 to 7 years. If criteria A, A1-A4, and C were met, criterion D was assessed; medical conditions reported by the parents were evaluated carefully for sufficiently explaining an eating disturbance causing problems with weight, growth, or nutrition. For example, although food allergies lead to some restriction of food intake, they were not considered sufficient to explain problems with weight, growth, or nutrition, as it is possible to consume substitutes for allergenic foods; criterion D was therefore considered as met.

The presence of any of the three drivers of food avoidance was not required to meet criteria for ARFID, as they are considered examples and not intended to be exhaustive (American Psychiatric Association, 2013; Bourne, Bryant-Waugh, Cook, & Mandy, 2020). A driver of food avoidance was considered present if the corresponding item was rated at least “sometimes” on a 5-point scale from “never” to “always” (Table 1). Drivers do not necessarily need to be present with all foods at all times, for example, there can be sensory-based avoidance of some foods, but not all or most foods. In this instance the parent might respond with “sometimes”, which is why we chose this response as the threshold to indicate evidence for a certain driver.

Initially, the ARFID screener was designed to assess both current and previous ARFID symptoms, in order to be able to determine point *and* lifetime prevalence of ARFID. Parents were therefore asked whether a problem was present *currently* or *previously*. During data analysis, we realized that the data basis to identify *previous* ARFID was insufficient. For example, as we had no indication of *when* certain problems were previously present, we could not ascertain that the ARFID criteria were met simultaneously at some point. Furthermore, almost all items were worded from a current perspective, providing a strong basis to evaluate current ARFID, but a less strong basis to evaluate previous ARFID. One item (assessing criterion A3) had to be excluded from the diagnostic algorithm, as the response options provided no indication of whether this problem was currently present or not (see documentation for item A3-c in Table S1).

Please note that in this study, “prevalence of ARFID” and “children with ARFID” refer to children screening positive for ARFID by meeting the diagnostic criteria as described above.

#### 2.2.2 Measurements to assess convergent validity

In addition to the questions screening for ARFID, parents filled in the child part of the *Behavioral Pediatric Feeding Assessment Scale* (BPFAS; William Crist et al., 1994; W. Crist & Napier-Phillips, 2001; with the permission of William Crist), which measures children’s restrictive-type eating behaviors, including problem behaviors related to mealtimes and nutritional intake. BPFAS items are rated on two scales: (1) a 5-point frequency scale from “never” to “always”; summing these ratings yields the *BPFAS Child Frequency Score*, and (2) a problem scale (“Is this a problem for you?” no=0, yes=1); summing these ratings yields the *BPFAS Child Problem Score*. Two BPFAS items relating to the food types eaten by the child were adapted slightly for the current study to enhance cultural relevance by adding more examples to the respective food group. Item 6 “Eats meat and/or fish” was changed to “Eats protein (e.g., meat, fish, eggs, beans, tofu)”, and item 18 “Eats starches (e.g. potato noodles)” was changed to “Eats starches (e.g., potatoes, rice, bread, pasta)”. The BPFAS has previously been shown to discriminate well between 2-to 7-year-old children with ARFID and a normative population (Dovey, Aldridge, Martin, Wilken, & Meyer, 2016). Children screening positive for ARFID were therefore expected to have significantly higher scores than children screening negative for ARFID on both BPFAS scales.

Parents also completed the following five subscales of the *Child Eating Behavior Questionnaire* (CEBQ; Carnell & Wardle, 2007; Wardle, Guthrie, Sanderson, & Rapoport, 2001): *Food Responsiveness, Food Fussiness, Satiety Responsiveness, Emotional Undereating, Emotional Overeating*. CEBQ items are rated on a 5-point frequency scale from “never” to “always”. Convergent validity would be evidenced if children screening positive for ARFID scored significantly *higher* on the scales Food Fussiness (selective eating, food neophobia), Satiety Responsiveness (feeling full quickly), and Emotional Undereating (eating less when having negative emotions), and if they scored significantly *lower* on the scales Food Responsiveness (having good appetite/being attracted by food, overeating) and Emotional Overeating (eating more when having negative emotions). The CEBQ scales *Enjoyment of Food* and *Slowness in Eating* could potentially have provided additional information on convergent validity, but were not included in the questionnaire to keep the questionnaire as concise as possible and to reduce the burden for participants.

Lastly, parents were asked to report their child’s height and weight at the time of answering the questionnaire. Although ARFID is not a low-weight disorder per se, children with ARFID are expected to be of lower weight and height on average, especially those meeting criterion A1 (weight loss or failure to grow/gain weight).

#### 2.2.3 Translation of the questionnaire into Japanese

The ARFID screener was originally developed in English. A Japanese native speaker (experienced child psychiatrist specialized in neurodevelopmental disorders and fluent in English) translated the ARFID screener, the BPFAS, and the CEBQ scales into Japanese. A native Swedish speaker with excellent knowledge of Japanese and English, working as a scientific English translator in our research center, translated back into English. Finally, the first author, the translator, and the back-translator discussed and resolved discrepancies.

### 2.3 Statistical Analyses

Point prevalence of ARFID was determined according to the algorithm described above and in Table 1. As the BPFAS and the CEBQ have not been used in Japan before, their psychometric properties were evaluated. Internal consistency of the total mean scores of the BPFAS and CEBQ scales was calculated using Cronbach’s alpha. Factorial validity of the CEBQ scales was examined using Principal Component Analysis (PCA) with orthogonal Varimax rotation. Extraction of factors was based on Eigenvalues >1 and scree plot inspections. Two-tailed Welch’s *t*-tests and Hedge’s *g* for effect size were used to investigate differences in BPFAS and CEBQ scale mean scores between children screening positive versus negative for ARFID. Hedge’s *g* is sample-bias correction for Cohen’s *d* (Lakens, 2013) and can therefore be interpreted similarly to Cohen’s *d* (suggested benchmarks: ≥0.2 small effect, ≥0.5 medium effect, ≥0.8 large effect (Cohen, 1988)). The significance level was set at 0.05. Using the parent-reported current height and weight of their child, height and body mass index (BMI) were evaluated using Japanese norm data collected in a national survey that included 14,000 0-6 year-olds and 695,600 6-17 year-olds (Kato, Murata, Kawano, Taniguchi, & Ohtake, 2004). The norm data are split by sex and month of age, and provide height in standard deviations (SDs) and BMI in percentile groups. Wilcoxon rank-sum tests were used to test group differences in height and BMI. Stata 16.1 was used for data analysis (StataCorp, 2019).

## 3. Results

### 3.1 Participant characteristics

Data were available from 3,746 children aged between 49 and 95 months (M=68.1, SD=11.0; 4 year-old: 26.6%, 5 year-old: 34.1%, 6 year-old: 28.0%, 7 year-old: 11.3%). 49.1% of the sample were female. Questionnaires were almost always filled in by mothers (98.1%), followed by fathers (1.8%), and in three cases by grandmothers. The mother was also indicated as a main caregiver for 98.1% of children (fathers: 60.1%; multiple answers were possible).

### 3.2 Prevalence of ARFID

The frequencies of responses to the single diagnostic criteria in the total cohort are shown in Table 1. A response to criterion A was missing in 14 children and in four children it was unclear whether criterion D was met. These 18 children were excluded from the analyses, leaving a total sample of 3,728 children. The point prevalence of ARFID was 1.3% (n=49). ARFID was slightly more common in girls (1.5%) than in boys (1.2%). The majority of children identified with ARFID displayed psychosocial impairment only (49.0%), more than a third (36.7%) showed physical impairment only, and 14.3% had both physical psychosocial and impairment. In other words, ARFID *with* physical impairment (51%, n=25) was equally common as ARFID *without* physical impairment (49%, n=24; Table 2). Criterion A1 (concerns about weight gain/growth) was met in almost all children with ARFID *with* physical impairment (96.0%, n=24), only a minority had nutritional deficiencies (criterion A2, 8%, n=2) or were dependent on nutritional supplements (criterion A3, 12% n=3). About a quarter (28%, n=7) of children with ARFID *with* physical impairment also experienced significant psychosocial impairment (criterion A4). Almost two thirds (63%, n=31) of *all* children with ARFID met criterion A4.

**Table 2.**
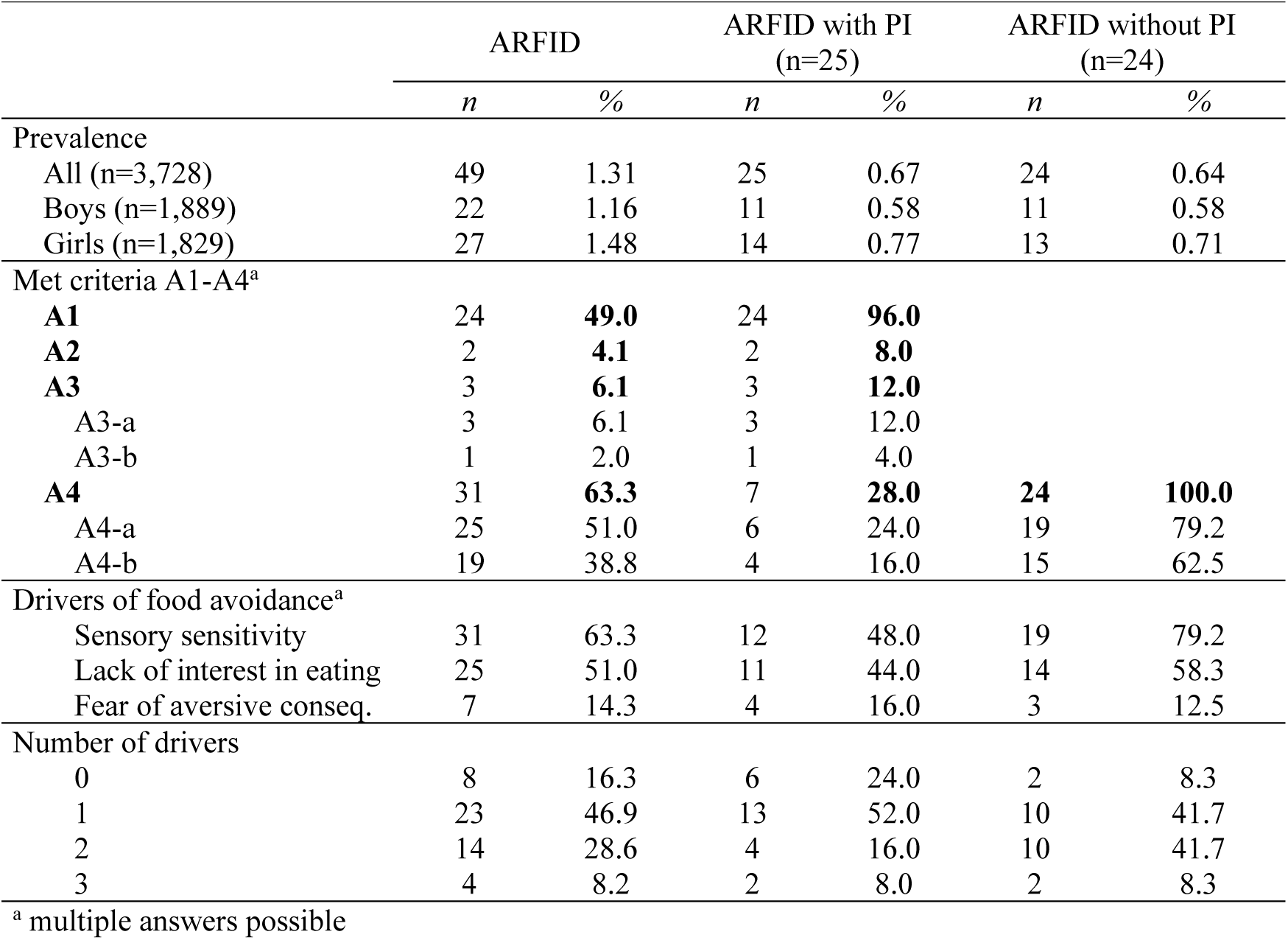
Prevalence of Avoidant/Restrictive Food Intake Disorder (ARFID) with and without physical impairment (PI) in Japanese children aged 4-7 years, met diagnostic criteria, and drivers of food avoidance

|  | ARFID |  | ARFID with PI<br>(n=25) |  | ARFID without PI<br>(n=24) |  |
| --- | --- | --- | --- | --- | --- | --- |
|  | *n* | % | *n* | % | *n* | % |
| Prevalence |  |  |  |  |  |  |
| All (n=3,728) | 49 | 1.31 | 25 | 0.67 | 24 | 0.64 |
| Boys (n=1,889) | 22 | 1.16 | 11 | 0.58 | 11 | 0.58 |
| Girls (n=1,829) | 27 | 1.48 | 14 | 0.77 | 13 | 0.71 |
| Met criteria A1-A4<sup>a</sup> |  |  |  |  |  |  |
| **A1** | 24 | **49.0** | 24 | **96.0** |  |  |
| **A2** | 2 | **4.1** | 2 | **8.0** |  |  |
| **A3** | 3 | **6.1** | 3 | **12.0** |  |  |
| A3-a | 3 | 6.1 | 3 | 12.0 |  |  |
| A3-b | 1 | 2.0 | 1 | 4.0 |  |  |
| **A4** | 31 | **63.3** | 7 | **28.0** | **24** | **100.0** |
| A4-a | 25 | 51.0 | 6 | 24.0 | 19 | 79.2 |
| A4-b | 19 | 38.8 | 4 | 16.0 | 15 | 62.5 |
| Drivers of food avoidance<sup>a</sup> |  |  |  |  |  |  |
| Sensory sensitivity | 31 | 63.3 | 12 | 48.0 | 19 | 79.2 |
| Lack of interest in eating | 25 | 51.0 | 11 | 44.0 | 14 | 58.3 |
| Fear of aversive conseq. | 7 | 14.3 | 4 | 16.0 | 3 | 12.5 |
| Number of drivers |  |  |  |  |  |  |
| 0 | 8 | 16.3 | 6 | 24.0 | 2 | 8.3 |
| 1 | 23 | 46.9 | 13 | 52.0 | 10 | 41.7 |
| 2 | 14 | 28.6 | 4 | 16.0 | 10 | 41.7 |
| 3 | 4 | 8.2 | 2 | 8.0 | 2 | 8.3 |

<sup>a</sup> multiple answers possible

### 3.3 Drivers of food avoidance

84% of children with ARFID reported at least one of the three drivers of food avoidance (boys: 82%, girls: 85%). Sensory sensitivity to food characteristics was the most common driver (63%), followed by lack of interest in eating (51%), while fear of aversive consequences of eating was much less frequent (14%; Table 2). Sensory sensitivity to food characteristics was somewhat more common in boys (77%, girls: 52%), while lack of interest in eating was somewhat more common in girls (59%, boys: 41%). While most children with ARFID (47%) showed evidence for only one of the drivers, 29% of the children had two drivers and 8% had three drivers.

### 3.4 Psychometric properties of the BPFAS and the CEBQ

Internal consistencies of the BPFAS scales were α=.80 for both the Child Frequency Score and the Child Problem Score; internal consistencies of the CEBQ scales ranged from α=.65 to α=.86 (Table S2). In the PCA for the five CEBQ scales, Eigenvalues suggested a five-factor solution accounting for 58% of the total variance, while the Screeplot suggested six factors accounting for 62% of the variance. We decided to explore item fits for the five-factor solution in order to enhance interpretability with the original CEBQ scales. The original scales were largely reproduced, except Food Responsiveness, where only two of the five items supposed to measure this scale had loadings larger than .30 (for details see Table S2). Due to lacking factorial validity, the Food Responsiveness scale was dropped from further analyses.

### 3.5 Convergent validity of the ARFID screener

Children with ARFID had significantly higher values than children screening negative for ARFID on the BPFAS Child Frequency Score (*g*_Hedge_=1.25) and the BPFAS Child Problem Score (*g*_Hedge_=1.50; Table 3). On average, parents of children with ARFID considered six different mealtime behaviors to be a problem, compared to two problematic mealtime behaviors in children screening negative for ARFID. As for the CEBQ scales, children with ARFID scored significantly higher than children without ARFID on Satiety Responsiveness (*g*_Hedge_=0.98), Food Fussiness (*g*_Hedge_=0.86), and Emotional Undereating (*g*_Hedge_=0.38). No significant differences were found for the scale Emotional Overeating. No significant differences in BPFAS and CEBQ scale scores were found between children with ARFID *with* physical impairment and children with ARFID but *without* physical impairment, however, due to the small groups, the power for these analyses was low, leading to wide confidence intervals.

**Table 3.** Comparison of Behavioral Pediatric Feeding Assessment Scale (BPFAS) scores and Child Eating Behavior Questionnaire (CEBQ) scores in children screening positive vs. negative for Avoidant/Restrictive Food Intake Disorder (ARFID), and in children screening positive for ARFID with vs without physical impairment (PI)

| ARFID vs. no ARFID | ARFID<br>(n=49) | no ARFID<br>(n=3,679) | Group comparison |  |  |
| --- | --- | --- | --- | --- | --- |
|  | *M (SD)* | *M (SD)* | *Welch's t (df)* | *p* | *Hedge's g (95% CI)<sup>a</sup>* |
| BPFAS |  |  |  |  |  |
| Child Frequency Score | 53.8 (11.1) | 43.2 (8.4) | 6.6 (48.8) | <.0001 | **1.25 (0.87, 1.62)** |
| Child Problem Score | 6.3 (4.2) | 2.2 (2.7) | 6.5 (44.5) | <.0001 | **1.50 (1.07, 1.93)** |
| CEBQ |  |  |  |  |  |
| Satiety Responsiveness | 3.2 (0.9) | 2.5 (0.7) | 5.2 (48.8) | <.0001 | **0.98 (0.63, 1.32)** |
| Food Fussiness | 3.4 (1.0) | 2.8 (0.8) | 4.9 (48.9) | <.0001 | **0.86 (0.53, 1.19)** |
| Emotional Undereating | 2.5 (0.8) | 2.3 (0.7) | 2.2 (48.9) | .034 | **0.38 (0.08, 0.67)** |
| Emotional Overeating | 1.5 (0.5) | 1.4 (0.4) | 1.1 (49.3) | .296 | 0.15 (-0.13, 0.44) |
| ARFID with vs. without PI | ARFID with<br>PI (n=25) | ARFID without<br>PI (n=24) | Group comparison |  |  |
|  | *M (SD)* | *M (SD)* | *Welch's t (df)* | *p* | *Hedge's g (95% CI)<sup>a</sup>* |
| BPFAS |  |  |  |  |  |
| Child Frequency Score | 51.2 (11.3) | 56.6 (10.4) | -1.7 (48.9) | .087 | -0.49 (-1.04, 0.07) |
| Child Problem Score | 5.5 (4.2) | 7.1 (4.2) | -1.3 (44.8) | .203 | -0.38 (-0.96, 0.20) |
| CEBQ |  |  |  |  |  |
| Satiety Responsiveness | 3.1 (1.0) | 3.3 (0.8) | -0.6 (45.8) | .553 | -0.17 (-0.72, 0.39) |
| Food Fussiness | 3.2 (1.0) | 3.7 (0.8) | -2.0 (47.4) | .050 | -0.56 (-1.12, 0.00) |
| Emotional Undereating | 2.3 (0.7) | 2.8 (0.9) | -2.0 (45.5) | .053 | -0.56 (-1.12, 0.01) |
| Emotional Overeating | 1.5 (0.5) | 1.6 (0.4) | -0.5 (49.0) | .618 | -0.14 (-0.69, 0.41) |

<sup>a</sup> Computed using Welch's formula to approximate the degrees of freedom (i.e., assuming unequal group variances).
Note. Possible score ranges: BPFAS Child Frequency Score: 25-125, BPFAS Child Problem Score: 0-25, CEBQ scales: 1-5

On average, children screening positive for ARFID were shorter in height and had noticeably lower BMI-for-age than children screening negative for ARFID, as indicated by descriptive numbers and Wilcoxon rank-sum tests (Table 4). However, this difference was mainly driven by the ARFID group *with* physical impairment: height and BMI were significantly lower in the ARFID group *with* physical impairment compared to children without ARFID (Z=2.17, p=.030; Z=2.92, p=.004, respectively). However, there were no significant differences in height and BMI between the ARFID group *without* physical impairment and children without ARFID (Z=1.09, p=.278; Z=0.27, p=.790, respectively; Table 4). The proportion of children with a BMI below the 10^th^ percentile (for sex and age) was 9.4% in the total sample, 13.6% in the ARFID group *without* physical impairment, but 36.4% in the ARFID group *with* physical impairment. Similarly, the proportion of children having a height of less than 2 SD below the mean (for sex and age) was 4.3% in the total sample, 4.4% in the ARFID group *without* physical impairment, but 26.1% in the ARFID group *with* physical impairment.

**Table 4.** Distribution of height in standard deviations (SDs) and body mass index (BMI) percentile groups in the total sample and in children screening positive for Avoidant/Restrictive Food Intake Disorder (ARFID) with and without physical impairment (PI) using Japanese norm data

| Height in SDs from the mean (M) | Total sample<br>(n=3,728) |  |  | ARFID<br>(n=49) |  |  | ARFID with PI<br>(n=25) |  |  | ARFID without PI<br>(n=24) |  |  |
| --- | --- | --- | --- | --- | --- | --- | --- | --- | --- | --- | --- | --- |
|  |  | *valid* | *cumul.* |  | *valid* | *cumul.* |  | *valid* | *cumul.* |  | *valid* | *cumul.* |
|  | *n* | % | % | *n* | % | % | *n* | % | % | *n* | % | % |
| < -3 SD | 26 | 0.8 | 0.8 | 3 | 6.5 | 6.5 | 2 | 8.7 | 8.7 | 1 | 4.4 | 4.4 |
| -3 to -2 SD<sup>a</sup> | 124 | 3.6 | 4.3 | 4 | 8.7 | 15.2 | 4 | 17.4 | 26.1 | 0 | 0.0 | 4.4 |
| -2 to -1.5 SD | 268 | 7.7 | 12.1 | 4 | 8.7 | 23.9 | 1 | 4.4 | 30.4 | 3 | 13.0 | 17.4 |
| -1.5 to -1 SD | 457 | 13.2 | 25.3 | 8 | 17.4 | 41.3 | 4 | 14.4 | 47.8 | 4 | 17.4 | 34.8 |
| -1 SD to M | 1309 | 37.8 | 63.1 | 14 | 30.4 | 71.7 | 6 | 26.1 | 73.9 | 8 | 34.8 | 69.6 |
| M to 1 SD | 955 | 27.6 | 90.7 | 10 | 21.7 | 93.5 | 4 | 17.4 | 91.3 | 6 | 26.1 | 95.7 |
| 1 to 1.5 SD | 203 | 5.9 | 96.6 | 1 | 2.2 | 95.7 | 0 | 4.4 | 91.3 | 1 | 4.4 | 100.0 |
| 1.5 to 2 SD | 87 | 2.5 | 99.1 | 1 | 2.2 | 97.8 | 1 | 0.0 | 95.7 | 0 | 0.0 | 100.0 |
| > 2 SD | 32 | 0.9 | 100.0 | 1 | 2.2 | 100.0 | 1 | 4.4 | 100.0 | 0 | 0.0 | 100.0 |
| Valid n (n missing) | 3,461<br>(267) |  |  | 46<br>(3) |  |  | 23<br>(2) |  |  | 23<br>(1) |  |  |
| Wilcoxon rank-sum tests: |  |  |  | ARFID vs. no ARFID |  |  | ARFID with PI vs.<br>no ARFID |  |  | ARFID without PI<br>vs. no ARFID |  |  |
|  |  |  |  | *Z* | *p* |  | *Z* | *p* |  | *Z* | *p* |  |
|  |  |  |  | 2.29 | .022 |  | 2.17 | .030 |  | 1.09 | .278 |  |

| BMI percentile group | Total sample<br>(n=3,728) |  |  | ARFID<br>(n=49) |  |  | ARFID with PI<br>(n=25) |  |  | ARFID without PI<br>(n=24) |  |  |
| --- | --- | --- | --- | --- | --- | --- | --- | --- | --- | --- | --- | --- |
|  |  | *valid* | *cumul.* |  | *valid* | *cumul.* |  | *valid* | *cumul.* |  | *valid* | *cumul.* |
|  | *n* | % | % | *n* | % | % | *n* | % | % | *n* | % | % |
| < 3rd | 118 | 3.4 | 3.4 | 4 | 9.1 | 9.1 | 4 | 18.2 | 18.2 | 0 | 0.0 | 0.0 |
| 3rd - 10th | 204 | 5.9 | 9.4 | 7 | 15.9 | 25.0 | 4 | 18.2 | 36.4 | 3 | 13.6 | 13.6 |
| 10th - 25th | 475 | 13.8 | 23.2 | 8 | 18.2 | 43.2 | 3 | 13.6 | 50.0 | 5 | 22.7 | 36.4 |
| 25th - 50th | 975 | 28.4 | 51.6 | 9 | 20.5 | 63.6 | 5 | 22.7 | 72.7 | 4 | 18.2 | 54.6 |
| 50th - 75th | 946 | 27.6 | 79.2 | 8 | 18.2 | 81.8 | 4 | 18.2 | 90.9 | 4 | 18.2 | 72.7 |
| 75th - 90th | 506 | 14.7 | 93.9 | 4 | 9.1 | 90.9 | 1 | 4.6 | 95.5 | 3 | 13.6 | 86.4 |
| 90th - 97th | 181 | 5.3 | 99.2 | 1 | 2.3 | 93.2 | 0 | 0.0 | 95.5 | 1 | 4.6 | 90.9 |
| 97th - 100th | 28 | 0.8 | 100.0 | 3 | 6.8 | 100.0 | 1 | 4.6 | 100.0 | 2 | 9.1 | 100.0 |
| Valid n (n missing) | 3,433<br>(295) |  |  | 44<br>(5) |  |  | 24<br>(3) |  |  | 22<br>(2) |  |  |
| Wilcoxon rank-sum tests: |  |  |  | ARFID vs. no ARFID |  |  | ARFID with PI vs.<br>no ARFID |  |  | ARFID without PI<br>vs. no ARFID |  |  |
|  |  |  |  | *Z* | *p* |  | *Z* | *p* |  | *Z* | *p* |  |
|  |  |  |  | 2.25 | .025 |  | 2.92 | .004 |  | 0.27 | .790 |  |

<sup>a</sup>The norm data were originally split into two categories here: [-3 to -2.5 SD] and [-2.5 to -2 SD]. However, none of the children with ARFID fell into the category [-3 to -2.5 SD], therefore the two categories were collapsed.

### 3.6 Modification of the screening tool – the ARFID-Brief Screener (ARFID-BS)

Based on the experience using the screening tool in the Japanese cohort and the data presented above, we have modified the original screener. The revised tool is called *ARFID-Brief Screener* (ARFID-BS, Supplement 2); it is intended to screen for ARFID in children from 2 years and up. Table S1 provides an item-by-item comparison of the original screener and the ARFID-BS, describes the modifications we made, and presents the diagnostic algorithm for the ARFID-BS. It is important to note that the ARFID-BS has not yet been validated; however, despite the modifications, the original screener and the ARFID-BS are very similar in the type of questions asked, in that the diagnostic algorithm maps closely onto the DSM-5 diagnostic criteria, as well as in the number of items included in the diagnostic algorithm. Although the findings from the present study cannot be directly transferred to the ARFID-BS, we do recommend future use of the ARFID-BS, and not of the original screener.

## 4. Discussion

The current study describes the development of a parent-reported screening tool for ARFID in children and its application in a large birth cohort of Japanese children aged 4 to 7 years. This is the largest prevalence study of ARFID for this age range, and the first to compare prevalence estimates for ARFID depending on the presence or absence of physical impairment.

Using the newly developed screener the point prevalence of children screening positive for ARFID was 1.3%. As ARFID prevalence might depend on age, this estimate is difficult to compare to previous studies. The use of different screening tools further reduces comparability across studies, as might also potential cultural differences. The only other study using a parent-reported instrument showed a considerably higher rate (15.5% in 5-10 year-olds) (Gonçalves et al., 2019); however, this rate seems disproportionately high in comparison to other prevalence estimates as discussed in the introduction. Studies using self-reports in older children (7-14 years) found higher (3.2-5.5% in Germany and Switzerland; Kurz et al., 2015; Schmidt et al., 2018) as well as lower estimates (0.3% in Taiwan; Chen et al., 2019). Over- or underreporting of symptoms are potential issues for both parent- and self-reports, but more research is needed to understand the extent to which they occur with respect to ARFID. For example, Schmidt et al. (2018) used the Eating Disorder in Youth-Questionnaire (EDY-Q; van Dyck et al., 2013) and found that only a quarter of children reporting ARFID symptoms and underweight problems in the EDY-Q were objectively underweight, while in Goncalves et al.’s study parental concern might have led to overreporting. That our study resulted in a significantly lower point prevalence than studies using the EDY-Q might be explained by our stricter criteria for ARFID; for example, as opposed to the EDY-Q, our ARFID screener evaluates exclusion criterion D and directly addresses actual restriction of food intake.

Whether physical consequences of avoidant and/or restrictive eating were required to be present had a large impact on ARFID prevalence: the number of children with ARFID doubled if physical consequences were not required, that is, if marked interferences with psychosocial functioning (criterion A4) was sufficient for screening positive for ARFID. This finding is difficult to compare, as previous epidemiological studies have not differentiated prevalence estimates by the presence or absence of physical consequences. However, the finding indicates that children with psychosocial impairment alone might make up a significant part of all cases with ARFID. Therefore, the clarification in DSM-5-TR that psychosocial impairment without physical impairment is sufficient for ARFID diagnosis seems highly appropriate. Apart from the impact on prevalence, future studies should also try to understand the association of physical consequences being present or absent with ARFID severity and other clinical characteristics (e.g., drivers of food avoidance). It is important to keep in mind that in this study, judgements regarding an interference with psychosocial functioning were made by parents, who might potentially overestimate the impact of their child’s eating pattern on their child’s stress and functioning by confusing it with their own stress and worry regarding their child’s eating. Especially in small children, criterion A4 can be difficult to evaluate, as parents often put a lot of effort into adjusting (family) life to their child’s demands around eating. More detailed guidelines on how to operationalize the A4 criterion in different age groups would be helpful for future research (for suggestions see (Harshman et al., 2021)).

Our results supported convergent validity of our ARFID screener with several measures. As expected, children screening positive for ARFID displayed more problems related to restrictive-type eating and nutritional intake as measured with the BPFAS. According to the CEBQ scales, children with ARFID were more often selective eaters and food neophobic (Food Fussiness), they were more likely to have lower levels of appetite and to eat less because of being sensitive to satiety (Satiety Responsiveness), and they tended to eat less when experiencing negative emotions (Emotional Undereating). While food fussiness is what we would expect to see mainly in children with sensory sensitivity, satiety responsiveness, and emotional undereating are largely associated with the lack-of-interest-in-eating-presentation of ARFID (Thomas et al., 2017). This was also found by He and colleagues (He, Zickgraf, Ellis, Lin, & Fan, 2021) who compared the Nine-Item ARFID screen (NIAS; Zickgraf & Ellis, 2018)—which assesses the three drivers of food avoidance with three items each— with the adult version of the CEBQ in Chinese adults. Furthermore, parent-reported height and weight data confirmed that children with ARFID *with* physical impairment (i.e., weight/height and nutrition problems) were in fact lighter in weight and shorter in height, while this was not the case for children with ARFID without physical impairment. This is in line with the results by Becker et al. (2019) who found that the average BMI of children with clinically diagnosed ARFID was in the normal range when allowing psychosocial impairment to be sufficient for ARFID diagnosis. Lastly, there were no significant differences in BPFAS and CEBQ scale scores between the ARFID group with and the ARFID group without physical impairment, indicating that also children with psychosocial impairment alone had highly problematic eating behaviors. Together, these data provide preliminary support for the newly developed screening tool used in this study. It differs from previously used instruments in two aspects. First, ARFID symptoms are parent-reported as opposed to self-reported. Second, previously used instruments focus strongly on the three known drivers of food avoidance, while consequences of avoidant/restrictive eating and exclusion criteria are included to a varying degree (e.g., EDY-Q, NIAS). In contrast, our ARFID screener is designed along the DSM-5 ARFID criteria and does not require the presence of any of the drivers of food avoidance.

As opposed to anorexia nervosa, where females are 3-10 times more often affected than males (Bulik et al., 2010; Hudson, Hiripi, Pope, & Kessler, 2007; Preti et al., 2009), the male-female ratio for ARFID in our sample was approximately 1:1. This is in line with previous epidemiological studies in children and adults (Chua et al., 2021; Hilbert, Zenger, Eichler, & Brähler, 2020; Kurz et al., 2015). Also studies from child and adolescent eating disorder programs, which are inherently female-based, found higher rates of males in patients with ARFID than in patients with anorexia nervosa (Fisher et al., 2014; Forman et al., 2014; Nicely, Lane-Loney, Masciulli, Hollenbeak, & Ornstein, 2014; Norris et al., 2014). Some studies of children referred to hospital-based pediatric feeding disorder programs even found boys to be in the majority (Sharp et al., 2020; Williams et al., 2015). The relatively much higher proportion of males in ARFID as compared with anorexia nervosa might be explained by two reasons. First, while overvaluation of body weight or shape and drive for thinness are part of the anorexia nervosa diagnostic criteria, they are exclusion criteria for the ARFID diagnosis. Such symptoms are known to be more common in females than in males (Anderson & Bulik, 2004; Nunez-Navarro et al., 2012), and this in itself might lead to a higher rate of diagnosable anorexia nervosa in females. Second, the ARFID symptoms *selective eating* and *sensory sensitivity* show strong associations with neurodevelopmental disorders (Ghanizadeh, 2011; Little, Dean, Tomchek, & Dunn, 2018; Robertson & Baron-Cohen, 2017; Sharp et al., 2013; Smith, Rogers, Blissett, & Ludlow, 2020), conditions that are generally present at higher rates in males than in females (Loomes, Hull, & Mandy, 2017; Polanczyk, de Lima, Horta, Biederman, & Rohde, 2007).

Among children with ARFID, sensory-based food avoidance was most common, while fear-based food avoidance was least common. This is concordant with previous studies in the general population (Kurz et al., 2015), but in contrast to studies of somewhat older children in partial hospitalization programs for eating disorders, where fear of aversive consequences has been found to be much more prevalent or even the most prevalent driver of food avoidance (Norris et al., 2018; Zickgraf, Lane-Loney, Essayli, & Ornstein, 2019). A reason for this might be that cases with acute onset—often triggered by a specific fear related to an aversive somatic experience—are overrepresented in clinical samples from intensive treatment programs, which might only represent a subgroup of children with ARFID in the general population. In addition, it can be speculated that fear-based food avoidance may be less prominent in young children, like the ones in our sample, as fear is usually associated with specific cognitions serving to maintain it. These cognitions require a certain level of cognitive development. In young children, traumatic experiences in eating might therefore more likely be reflected in lowered interest and sensory-based rejection of foods.

Having more than one driver of food avoidance was very common (36.8%), although less common than in a clinical study of pediatric patients with ARFID (64.4%), which explicitly aimed to investigate the overlap between drivers of food avoidance using somewhat more detailed questions (Reilly, Brown, Gray, Kaye, & Menzel, 2019). These observations contradict the notion of mutually exclusive ARFID subtypes, but are in line with the view that several drivers of food avoidance often occur together, with more drivers potentially exacerbating the condition (Thomas et al., 2017). A significant proportion (17.9%) did not show evidence for any of the three drivers. This could reflect the relatively simplistic measurement of possible drivers in this study with one question each, and/or indicate the existence of other potential and yet unknown reasons for food avoidance. In order to acknowledge the hypothesis that the three known drivers of food avoidance are not exhaustive (Bourne et al., 2020), our screening tool does not include questions relating to the drivers of food avoidance into the algorithm to identify children with potential ARFID, which is different from other tools (e.g., the EDY-Q). Future clinical studies should test if further drivers of food avoidance can be identified; if not, the three known drivers should be included into algorithms to identify ARFID.

This study has several limitations. First, the ARFID prevalence estimates reported in this study have to be considered preliminary until future studies provide evidence for the diagnostic validity of the ARFID-BS using clinically ascertained ARFID diagnoses. However, we found promising initial evidence of convergent validity with a range of relevant measures assessing restrictive-type eating as well as with weight and height. It should also be pointed out that the ARFID screening items used here are closely mapped onto the DSM-5 ARFID criteria and they therefore likely have high content validity. This method (i.e., to create screening items that almost verbatim reflect the DSM diagnostic criteria) has been used previously for a parent-reported questionnaire on neurodevelopmental disorders and led to high criterion validity for diagnoses of ASD and ADHD (Mårland et al., 2017). Second, the question applied to assess criterion D only partially reflected the exclusion criterion, that is, it did not link any concurrent medical condition to the food avoidance/restriction as such and it did not address other mental disorders (even though mental disorders other than autism spectrum disorder and Attention-Deficit/Hyperactivity Disorder are very infrequently diagnosed in this young age group). This might have led to an underestimation of children with medical or mental conditions that directly account for their eating disturbance, hence we possibly slightly overestimated the prevalence of ARFID. In the revised version of the screener, the ARFID-BS, we have adjusted this question to more accurately reflect DSM-5 criterion D (Supplement 2). Third, height and weight data were parent-reported and not objectively measured, which can be a source for (social desirability) bias, for instance, as parents tend to report higher weights for underweight children and lower weights for overweight children (van Leeuwen, van Middelkoop, Paulis, Bindels, & Koes, 2019; Wright, Glanz, Colburn, Robson, & Saelens, 2018). Considering that children with ARFID in our sample were more often underweight, we could have underestimated the difference in BMI between children with and without ARFID. Lastly, although the initially enrolled JECS cohort is representative for the Japanese population (Michikawa et al., 2018), response rates have declined with increasing time of follow-up, in line with other large longitudinal cohort studies on child development in Europe (Fraser et al., 2013; Olsen et al., 2019); the response rate in the current study was 56.5%. Mothers who dropped out during the first year of the JECS and their children seem to be less healthy on average (Kigawa et al., 2019). Our attrition analysis furthermore showed that mothers still participating in the JECS, but not responding to our questionnaire, might be slightly less well-functioning than responders, as indicated by lower socio-economic status and a few maternal health variables. Hence, it is possible that the children in our sample are slightly healthier than the average Japanese child population and that we therefore somewhat underestimated the prevalence of ARFID.

In summary, this study contributes to the small body of literature on the prevalence of ARFID in the general population and addresses the lack of parent-reported screening tools for ARFID. We present a newly developed ARFID screener and initial evidence for its validity. We also introduce a slightly modified screener, which we recommend to use in future validation studies of ARFID screeners. Such studies are urgently needed as valid screening instruments are an essential prerequisite for identifying individuals with ARFID, not only in large-scale epidemiological studies, but also in clinical practice. In a large birth cohort of 4-7-year-old Japanese children, 1.3% screened positive for ARFID. This demonstrates that ARFID is neither very common nor very rare. About half of the children identified with ARFID did not show evidence of physical impairment from avoidant and/or restrictive eating. This indicates that children with psychosocial impairment alone might make up a significant part of all cases with ARFID and testifies to the importance of including this aspect into screening instruments. More detailed guidelines on how to operationalize the assessment of psychosocial impairment in different age groups will be needed.

## Data Availability

Data are unsuitable for public deposition due to ethical restrictions and legal framework of Japan. It is prohibited by the Act on the Protection of Personal Information (Act No. 57 of 30 May 2003, amendment on 9 September 2015) to publicly deposit the data containing personal information. Ethical Guidelines for Medical and Health Research Involving Human Subjects enforced by the Japan Ministry of Education, Culture, Sports, Science and Technology and the Ministry of Health, Labour and Welfare also restricts the open sharing of the epidemiologic data. All inquiries about access to data should be sent to. The person responsible for handling enquiries sent to this e-mail address is Dr Shoji F. Nakayama, JECS Programme Office, National Institute for Environmental Studies.

## Abbreviations

ARFID: Avoidant/Restrictive Food Intake Disorder
ARFID-BS: ARFID-Brief Screener
BPFAS: Behavioral Pediatric Feeding Assessment Scale
CEBQ: Child Eating Behavior Questionnaire
EDY-Q: Eating Disorder in Youth-Questionnaire
JECS: Japan Environment and Children’s Study

## Acknowledgements

We would like to express our gratitude to all of the JECS study participants in the Kochi cohort and to the staff members at Kochi Regional Centre, who sent out and collected the questionnaires. Furthermore, we gratefully thank Theo Gillberg for back-translating the Early Eating Behavior Questionnaire (EEBQ) from Japanese. In addition, we thank Nadia Micali, who consulted us on the EEBQ in the early stages of its development.

## Author Contributions

L.D., M.R. and C.G. designed the study.

L.D., M.R. and C.G. obtained funding for this study.

L.D. performed the data analysis and drafted the manuscript.

K.Y.-L. coordinated the data collection.

K.Y.-L., M.E., M.F., N.S. and Y.H. provided administrative support for the data collection.

L.D., R.B.-W., M.R. and C.G. contributed to the interpretation of results.

R.B.-W., M.R. and C.G. supervised the study and provided clinical expertise.

All authors critically revised the manuscript for important intellectual content and approved the final manuscript.

## Previous peer-review

A previous version of this study has been peer-reviewed by the journal *European Eating Disorders Review*. We have changed the paper significantly and addressed the reviewers’ comments to the extent it was possible.

## Footnotes

1 At the start of JECS, 7,094 children were registered in the Kochi cohort. At the time our questionnaire was sent out, 6,633 of these were still participating in the JECS.

## References

American Psychiatric Association. (2013). Diagnostic and Statistical Manual of Mental Disorders, Fifth Edition. Arlington, VA: American Psychiatric Association.

Anderson, C. B., & Bulik, C. M. (2004). Gender differences in compensatory behaviors, weight and shape salience, and drive for thinness. Eating Behaviors, 5(1), 1–11. doi:https://doi.org/10.1016/j.eatbeh.2003.07.001

Becker, K. R., Keshishian, A. C., Liebman, R. E., Coniglio, K. A., Wang, S. B., Franko, D. L., … Thomas, J. J. (2019). Impact of expanded diagnostic criteria for avoidant/restrictive food intake disorder on clinical comparisons with anorexia nervosa. International Journal of Eating Disorders, 52(3), 230–238. doi:10.1002/eat.22988

Bourne, L., Bryant-Waugh, R., Cook, J., & Mandy, W. (2020). Avoidant/restrictive food intake disorder: A systematic scoping review of the current literature. Psychiatry Research, 288, 112961. doi:10.1016/j.psychres.2020.112961

Bulik, C. M., Thornton, L. M., Root, T. L., Pisetsky, E. M., Lichtenstein, P., & Pedersen, N. L. (2010). Understanding the relation between anorexia nervosa and bulimia nervosa in a Swedish national twin sample. Biological Psychiatry, 67(1), 71–77. doi:10.1016/j.biopsych.2009.08.010

Carnell, S., & Wardle, J. (2007). Measuring behavioural susceptibility to obesity: validation of the child eating behaviour questionnaire. Appetite, 48(1), 104–113. doi:10.1016/j.appet.2006.07.075

Chen, Y. L., Chen, W. J., Lin, K. C., Shen, L. J., & Gau, S. S. (2019). Prevalence of DSM-5 mental disorders in a nationally representative sample of children in Taiwan: methodology and main findings. Epidemiol Psychiatr Sci, 29, e15. doi:10.1017/s2045796018000793

Chua, S. N., Fitzsimmons-Craft, E. E., Austin, S. B., Wilfley, D. E., & Taylor, C. B. (2021). Estimated prevalence of eating disorders in Singapore. International Journal of Eating Disorders, 54(1), 7–18. doi:10.1002/eat.23440

Cohen, J. (1988). Statistical power analysis for the behavioral sciences. New York, NY: Routledge Academic.

Crist, W., McDonnell, P., Beck, M., Gillespie, C. T., Barrett, P., & Mathews, J. (1994). Behavior at Mealtimes and the Young Child with Cystic Fibrosis. Journal of Developmental and Behavioral Pediatrics, 15(3), 157–161. Retrieved from http://journals.lww.com/jrnldbp/Fulltext/1994/06000/Behavior_at_Mealtimes_and_the_Young_Child_with.1.aspx

Crist, W., & Napier-Phillips, A. (2001). Mealtime behaviors of young children: a comparison of normative and clinical data. Journal of Developmental and Behavioral Pediatrics, 22(5), 279–286.

Dinkler, L., & Bryant-Waugh, R. (2021). Assessment of avoidant restrictive food intake disorder, pica and rumination disorder: interview and questionnaire measures. Current Opinion in Psychiatry. doi:10.1097/yco.0000000000000736

Dovey, T. M., Aldridge, V. K., Martin, C. I., Wilken, M., & Meyer, C. (2016). Screening Avoidant/Restrictive Food Intake Disorder (ARFID) in children: Outcomes from utilitarian versus specialist psychometrics. Eating Behaviors, 23, 162-167. doi:http://dx.doi.org/10.1016/j.eatbeh.2016.10.004

Eddy, K. T., Harshman, S. G., Becker, K. R., Bern, E., Bryant-Waugh, R., Hilbert, A., … Thomas, J. J. (2019). Radcliffe ARFID Workgroup: Toward operationalization of research diagnostic criteria and directions for the field. International Journal of Eating Disorders, 52(4), 361–366. doi:10.1002/eat.23042

Fisher, M. M., Rosen, D. S., Ornstein, R. M., Mammel, K. A., Katzman, D. K., Rome, E. S., … Walsh, B. T. (2014). Characteristics of avoidant/restrictive food intake disorder in children and adolescents: a “new disorder” in DSM-5. Journal of Adolescent Health, 55(1), 49–52. doi:10.1016/j.jadohealth.2013.11.013

Fitzsimmons-Craft, E. E., Balantekin, K. N., Graham, A. K., Smolar, L., Park, D., Mysko, C., … Wilfley, D. E. (2019). Results of disseminating an online screen for eating disorders across the U.S.: Reach, respondent characteristics, and unmet treatment need. International Journal of Eating Disorders, 52(6), 721–729. doi:https://doi.org/10.1002/eat.23043

Forman, S. F., McKenzie, N., Hehn, R., Monge, M. C., Kapphahn, C. J., Mammel, K. A., … Woods, E. R. (2014). Predictors of outcome at 1 year in adolescents with DSM-5 restrictive eating disorders: report of the national eating disorders quality improvement collaborative. Journal of Adolescent Health, 55(6), 750–756. doi:10.1016/j.jadohealth.2014.06.014

Fraser, A., Macdonald-Wallis, C., Tilling, K., Boyd, A., Golding, J., Davey Smith, G., … Lawlor, D. A. (2013). Cohort Profile: the Avon Longitudinal Study of Parents and Children: ALSPAC mothers cohort. International Journal of Epidemiology, 42(1), 97–110. doi:10.1093/ije/dys066

Ghanizadeh, A. (2011). Sensory Processing Problems in Children with ADHD, a Systematic Review. Psychiatry Investigation, 8(2), 89–94. doi:10.4306/pi.2011.8.2.89

Gonçalves, S., Vieira, A. I., Machado, B. C., Costa, R., Pinheiro, J., & Conceiçao, E. (2019). Avoidant/restrictive food intake disorder symptoms in children: Associations with child and family variables. Children’s Health Care, 48(3), 301–313. doi:10.1080/02739615.2018.1532796

Harshman, S. G., Jo, J., Kuhnle, M., Hauser, K., Murray, H. B., Becker, K. R., … Thomas, J. J. (2021). A Moving Target: How We Define Avoidant/Restrictive Food Intake Disorder Can Double Its Prevalence. Journal of Clinical Psychiatry, 82(5). doi:10.4088/JCP.20m13831

Hay, P., Mitchison, D., Collado, A. E. L., Gonzalez-Chica, D. A., Stocks, N., & Touyz, S. (2017). Burden and health-related quality of life of eating disorders, including Avoidant/Restrictive Food Intake Disorder (ARFID), in the Australian population. J Eat Disord, 5, 21. doi:10.1186/s40337-017-0149-z

He, J., Zickgraf, H. F., Ellis, J. M., Lin, Z., & Fan, X. (2021). Chinese Version of the Nine Item ARFID Screen: Psychometric Properties and Cross-Cultural Measurement Invariance. Assessment, 28(2), 537–550. doi:10.1177/1073191120936359

Hilbert, A., Zenger, M., Eichler, J., & Brahler, E. (2021). Psychometric evaluation of the Eating Disorders in Youth-Questionnaire when used in adults: Prevalence estimates for symptoms of avoidant/restrictive food intake disorder and population norms. International Journal of Eating Disorders, 54(3), 399–408. doi:10.1002/eat.23424

Hilbert, A., Zenger, M., Eichler, J., & Brähler, E. (2020). Psychometric evaluation of the Eating Disorders in Youth-Questionnaire when used in adults: Prevalence estimates for symptoms of avoidant/restrictive food intake disorder and population norms. Int J Eat Disord. doi:10.1002/eat.23424

Hudson, J. I., Hiripi, E., Pope, H. G., Jr., & Kessler, R. C. (2007). The prevalence and correlates of eating disorders in the National Comorbidity Survey Replication. Biological Psychiatry, 61(3), 348–358. doi:10.1016/j.biopsych.2006.03.040

Kato, N., Murata, M., Kawano, M., Taniguchi, T., & Ohtake, T. (2004). Growth standard for children from 0 up to 18 years of age (in Japanese). Shonihokenkenkyu, 63(5), 345–348.

Kawamoto, T., Nitta, H., Murata, K., Toda, E., Tsukamoto, N., Hasegawa, M., … Satoh, H. (2014). Rationale and study design of the Japan environment and children’s study (JECS). BMC Public Health, 14, 25. doi:10.1186/1471-2458-14-25

Kigawa, M., Tsuchida, A., Matsumura, K., Takamori, A., Ito, M., Tanaka, T., … Inadera, H. (2019). Factors of non-responsive or lost-to-follow-up Japanese mothers during the first year post partum following the Japan Environment and Children’s Study: a longitudinal cohort study. BMJ Open, 9(11), e031222. doi:10.1136/bmjopen-2019-031222

Kurz, S., van Dyck, Z., Dremmel, D., Munsch, S., & Hilbert, A. (2015). Early-onset restrictive eating disturbances in primary school boys and girls. European Child and Adolescent Psychiatry, 24(7), 779–785. doi:10.1007/s00787-014-0622-z

Lakens, D. (2013). Calculating and reporting effect sizes to facilitate cumulative science: a practical primer for t-tests and ANOVAs. Frontiers in Psychology, 4, 863. doi:10.3389/fpsyg.2013.00863

Little, L. M., Dean, E., Tomchek, S., & Dunn, W. (2018). Sensory Processing Patterns in Autism, Attention Deficit Hyperactivity Disorder, and Typical Development. Physical & Occupational Therapy in Pediatrics, 38(3), 243–254. doi:10.1080/01942638.2017.1390809

Loomes, R., Hull, L., & Mandy, W. P. L. (2017). What Is the Male-to-Female Ratio in Autism Spectrum Disorder? A Systematic Review and Meta-Analysis. Journal of the American Academy of Child and Adolescent Psychiatry, 56(6), 466–474. doi:10.1016/j.jaac.2017.03.013

Michikawa, T., Nitta, H., Nakayama, S. F., Yamazaki, S., Isobe, T., Tamura, K., … Kawamoto, T. (2018). Baseline Profile of Participants in the Japan Environment and Children’s Study (JECS). Journal of Epidemiology, 28(2), 99–104. doi:10.2188/jea.JE20170018

Mårland, C., Lichtenstein, P., Degl’Innocenti, A., Larson, T., Råstam, M., Anckarsäter, H., … Lundström, S. (2017). The Autism-Tics, ADHD and other Comorbidities inventory (A-TAC): previous and predictive validity. BMC Psychiatry, 17(1), 403. doi:10.1186/s12888-017-1563-0

Nicely, T. A., Lane-Loney, S., Masciulli, E., Hollenbeak, C. S., & Ornstein, R. M. (2014). Prevalence and characteristics of avoidant/restrictive food intake disorder in a cohort of young patients in day treatment for eating disorders. J Eat Disord, 2(1), 21. doi:10.1186/s40337-014-0021-3

Norris, M. L., Robinson, A., Obeid, N., Harrison, M., Spettigue, W., & Henderson, K. (2014). Exploring avoidant/restrictive food intake disorder in eating disordered patients: a descriptive study. International Journal of Eating Disorders, 47(5), 495–499. doi:10.1002/eat.22217

Norris, M. L., Spettigue, W., Hammond, N. G., Katzman, D. K., Zucker, N., Yelle, K., … Obeid, N. (2018). Building evidence for the use of descriptive subtypes in youth with avoidant restrictive food intake disorder. International Journal of Eating Disorders, 51(2), 170–173. doi:10.1002/eat.22814

Nunez-Navarro, A., Aguera, Z., Krug, I., Jimenez-Murcia, S., Sanchez, I., Araguz, N., … Fernandez-Aranda, F. (2012). Do men with eating disorders differ from women in clinics, psychopathology and personality? European Eating Disorders Review, 20(1), 23–31. doi:10.1002/erv.1146

Olsen, E. M., Rask, C. U., Elberling, H., Jeppesen, P., Clemmensen, L., Munkholm, A., … Skovgaard, A. M. (2019). Cohort Profile: The Copenhagen Child Cohort Study (CCC2000). International Journal of Epidemiology. doi:10.1093/ije/dyz256

Polanczyk, G., de Lima, M. S., Horta, B. L., Biederman, J., & Rohde, L. A. (2007). The worldwide prevalence of ADHD: a systematic review and metaregression analysis. American Journal of Psychiatry, 164(6), 942–948. doi:10.1176/ajp.2007.164.6.942

Preti, A., Girolamo, G., Vilagut, G., Alonso, J., Graaf, R., Bruffaerts, R., … Morosini, P. (2009). The epidemiology of eating disorders in six European countries: results of the ESEMeD-WMH project. Journal of Psychiatric Research, 43(14), 1125–1132. doi:10.1016/j.jpsychires.2009.04.003

Reilly, E. E., Brown, T. A., Gray, E. K., Kaye, W. H., & Menzel, J. E. (2019). Exploring the cooccurrence of behavioural phenotypes for avoidant/restrictive food intake disorder in a partial hospitalization sample. European Eating Disorders Review, 27(4), 429–435. doi:10.1002/erv.2670

Robertson, C. E., & Baron-Cohen, S. (2017). Sensory perception in autism. Nature Reviews: Neuroscience, 18(11), 671–684. doi:10.1038/nrn.2017.112

Schmidt, R., Vogel, M., Hiemisch, A., Kiess, W., & Hilbert, A. (2018). Pathological and non-pathological variants of restrictive eating behaviors in middle childhood: A latent class analysis. Appetite, 127, 257–265. doi:10.1016/j.appet.2018.04.030

Sharp, W. G., Berry, R. C., McCracken, C., Nuhu, N. N., Marvel, E., Saulnier, C. A., … Jaquess, D. L. (2013). Feeding problems and nutrient intake in children with autism spectrum disorders: a meta-analysis and comprehensive review of the literature. Journal of Autism and Developmental Disorders, 43(9), 2159–2173. doi:10.1007/s10803-013-1771-5

Sharp, W. G., Volkert, V. M., Stubbs, K. H., Berry, R. C., Clark, M. C., Bettermann, E. L., … Scahill, L. (2020). Intensive Multidisciplinary Intervention for Young Children with Feeding Tube Dependence and Chronic Food Refusal: An Electronic Health Record Review. Journal of Pediatrics, 223, 73–80 e72. doi:10.1016/j.jpeds.2020.04.034

Smith, B., Rogers, S. L., Blissett, J., & Ludlow, A. K. (2020). The relationship between sensory sensitivity, food fussiness and food preferences in children with neurodevelopmental disorders. Appetite, 150, 104643. doi:10.1016/j.appet.2020.104643

StataCorp. (2019). Stata Statistical Software: Release 16. College Station, TX: StataCorp LLC.

Thomas, J. J., Lawson, E. A., Micali, N., Misra, M., Deckersbach, T., & Eddy, K. T. (2017). Avoidant/Restrictive Food Intake Disorder: a Three-Dimensional Model of Neurobiology with Implications for Etiology and Treatment. Curr Psychiatry Rep, 19(8), 54. doi:10.1007/s11920-017-0795-5

van Dyck, Z., Bellwald Kurz, S., Dremmel, D., Munsch, S., & Hilbert, A. (2013). Essprobleme im Kindesalter. Screening in der allgemeinen Bevölkerung [Eating disorders in childhood and adolescence]. Zeitschrift für Gesundheitspsychologie, 21(2), 91–100. doi:10.1026/0943-8149/a000091

van Leeuwen, J., van Middelkoop, M., Paulis, W. D., Bindels, P. J. E., & Koes, B. W. (2019). General practitioners cannot rely on reported weight and height of children. Primary Health Care Research & Development, 20, e14–e14. doi:10.1017/S1463423618000713

Wardle, J., Guthrie, C. A., Sanderson, S., & Rapoport, L. (2001). Development of the Children’s Eating Behaviour Questionnaire. Journal of Child Psychology and Psychiatry and Allied Disciplines, 42(7), 963–970.

Williams, K. E., Hendy, H. M., Field, D. G., Belousov, Y., Riegel, K., & Harclerode, W. (2015). Implications of Avoidant/Restrictive Food Intake Disorder (ARFID) on Children with Feeding Problems. Children’s Health Care, 44(4), 307–321. doi:10.1080/02739615.2014.921789

World Health Organization. (2018). International Classification of Diseases for Mortality and Morbidity Statistics, Eleventh Revision. Retrieved from https://icd.who.int/browse11/l-m/en

Wright, D. R., Glanz, K., Colburn, T., Robson, S. M., & Saelens, B. E. (2018). The accuracy of parent-reported height and weight for 6-12 year old U.S. children. BMC Pediatrics, 18(1), 52–52. doi:10.1186/s12887-018-1042-x

Zickgraf, H. F., & Ellis, J. M. (2018). Initial validation of the Nine Item Avoidant/Restrictive Food Intake disorder screen (NIAS): A measure of three restrictive eating patterns. Appetite, 123, 32–42. doi:10.1016/j.appet.2017.11.111

Zickgraf, H. F., Lane-Loney, S., Essayli, J. H., & Ornstein, R. M. (2019). Further support for diagnostically meaningful ARFID symptom presentations in an adolescent medicine partial hospitalization program. International Journal of Eating Disorders, 52(4), 402–409. doi:10.1002/eat.23016

