## Supplementary Material for "Development of a parent-reported screening tool for avoidant/restrictive food intake disorder (ARFID): Initial validation and prevalence in 4-7-year-old Japanese children"

**Supplement 1:** Comparison of responders and non-responders

Comparing responders (56.5%) and non-responders to the questionnaire in the Kochi cohort (N=6,633) we found that responding mothers were more highly educated, had a higher annual household income, and smoked less often during pregnancy. With regard to maternal health variables, few differences were found, except that responding mothers were less often overweight prior to pregnancy and experienced less psychological distress during pregnancy. No differences emerged for diagnoses of depression, anxiety disorders, and schizophrenia, autistic traits, overeating and measures to reduce weight in the year prior to pregnancy. Children of responding mothers were slightly less often born preterm, however, children's Apgar scores did not differ between responders and non-responders.

| | **Responders (n=3,746)** | **Non-responders (n=2,887)** | |
|---|---|---|---|
| **General characteristics** | % | % | p |
| Maternal age at study entry (during trimester 1) | | | **<.001** |
| Total, mean (SD) | 31.2 (4.7) | 30.3 (5.1) | |
| <25 | 7.3 | 13.6 | |
| 25-29 | 29.9 | 30.4 | |
| 30-34 | 37.1 | 33.7 | |
| ≥35 | 25.7 | 22.3 | |
| Maternal education, years | | | **<.001** |
| <10 | 3.1 | 5.9 | |
| 10-12 | 23.2 | 30.1 | |
| 13-16 | 72.0 | 63.0 | |
| ≥17 | 1.7 | 1.0 | |
| Annual household income, million Japanese Yen | | | **<.001** |
| < 2 | 7.5 | 11.0 | |
| 2 to <4 | 33.6 | 39.3 | |
| 4 to <6 | 32.9 | 29.0 | |
| 6 to <8 | 17.9 | 13.6 | |
| 8 to <10 | 5.2 | 4.8 | |
| ≥10 | 2.8 | 2.3 | |
| Any person smoking in the household | 19.9 | 28.5 | **<.001** |
| **Child/delivery characteristics** | % | % | p |
| Sex, % female | 49.1 | 49.2 | .960 |
| Gestational age at birth, weeks | | | .100 |
| Total, mean (SD) | 39.1 (1.6) | 39.0 (1.7) | |
| Preterm births (<37) | 5.3 | 6.5 | |
| Term births (37-41) | 94.5 | 93.4 | |
| Postterm births (≥42) | 0.1 | 0.1 | |
| Birth weight in g, mean (SD) | 2987.2 (413.8) | 2976.1 (428.9) | .292 |
| Multiple births | 1.3 | 1.6 | .345 |
| Apgar score < 7 at 1 minute after birth | 2.5 | 2.7 | .752 |
| Apgar score < 7 at 5 minutes after birth | 0.8 | 1.0 | .493 |
| **Maternal health** | % | % | p |
| BMI prior to pregnancy, $kg/m^2$ | | | **.005** |
| Total, mean (SD) | 21.09 (3.06) | 21.35 (3.33) | |
| <18.5 | 15.6 | 15.6 | |
| 18.5-24.9 | 75.2 | 72.7 | |
| ≥25.0 | 9.2 | 11.6 | |
| Maternal smoking during pregnancy | | | **<.001** |
| Never smoked | 64.4 | 57.5 | |
| Stopped smoking before pregnancy was noticed | 20.7 | 20.2 | |
| Stopped smoking when pregnancy was noticed | 10.6 | 14.9 | |
| Still smoking | 4.3 | 7.4 | |

| Maternal health | Responders (n=3,746) % | Non-responders (n=2,887) % | p |
|---|---|---|---|
| Maternal alcohol consumption during pregnancy | | | .578 |
| Never drank | 31.8 | 31.9 | |
| Stopped drinking before pregnancy was noticed | 18.3 | 17.0 | |
| Stopped drinking when pregnancy was noticed | 47.1 | 48.1 | |
| Still drinking | 2.8 | 3.0 | |
| Mental disorders (current or previous) | | | |
| Depression | 2.3 | 2.8 | .207 |
| Anxiety disorders | 2.3 | 2.4 | .806 |
| Schizophrenia | 0.05 | 0.04 | 1 |
| Health-related quality of life (SF-8) | | | |
| Trimester 1 | | | |
| Physical composite score, mean (SD) | 45.2 (7.4) | 45.5 (7.2) | .154 |
| Mental composite score, mean (SD) | 46.7 (7.3) | 46.5 (7.3) | .302 |
| Trimester 2 | | | |
| Physical composite score, mean (SD) | 46.0 (6.4) | 45.9 (6.4) | .391 |
| Mental composite score, mean (SD) | 49.6 (6.2) | 49.3 (6.3) | **.048** |
| At child age 2.5 years | | | |
| Physical composite score, mean (SD) | 49.5 (6.3) | 49.5 (6.4) | .844 |
| Mental composite score, mean (SD) | 48.3 (6.4) | 48.0 (6.5) | .112 |
| Psychological distress/mental health (K-6 ≥ 13) | | | |
| Trimester 1 | 2.4 | 3.2 | **.046** |
| Trimester 2 | 2.3 | 3.5 | **.008** |
| At child age 2.5 years | 2.3 | 2.0 | .585 |
| Autism Quotient (AQ-10) ≥ 6 | 7.0 | 6.3 | .272 |
| Are you a person who tends to overeat? | | | |
| Trimester 1 | 29.9 | 30.1 | .828 |
| Trimester 2 | 24.9 | 23.9 | .338 |
| Measures to reduce weight as a teenager | | | |
| Reduce the amount of food to two-thirds or less of the regular amount | 26.7 | 26.8 | 1 |
| Reduce snacks between meals and in the evenings | 49.8 | 50.9 | .384 |
| Eat particular foods (diet foods) | 20.3 | 19.4 | .334 |
| Use (pharmaceutical) drug | 5.6 | 5.6 | .957 |
| Purging | 6.0 | 5.6 | .489 |
| Smoke cigarettes | 6.3 | 8.0 | **.011** |
| Exercise | 47.0 | 44.1 | **.025** |
| Any measure to reduce weight | 71.1 | 71.2 | .956 |
| Measures to reduce weight in the year prior to pregnancy | | | |
| Reduce the amount of food to two-thirds or less of the regular amount | 12.2 | 13.3 | .217 |
| Reduce snacks between meals and in the evenings | 36.3 | 37.6 | .279 |
| Eat particular foods (diet foods) | 9.8 | 9.9 | .901 |
| Use (pharmaceutical) drug | 2.0 | 2.0 | 1 |
| Purging | 2.1 | 2.0 | .792 |
| Smoke cigarettes | 3.5 | 5.4 | **<.001** |
| Exercise | 32.5 | 32.1 | .749 |
| Any measure to reduce weight | 55.7 | 56.9 | .316 |

Note. Fisher's exact test was used for association tests, except when mean (SD) are given—in these cases Welch's t-test was used. Associations significant at the .05 level are printed in bold. No correction for multiple testing was applied.

**Supplement 2:** ARFID-Brief Screener (ARFID-BS)

Today's date (yymmdd): ☐☐☐☐☐☐

Participant code (filled in by research staff): ☐☐☐☐

### Questionnaire on eating problems in childhood

This questionnaire contains questions that describe certain eating problems in childhood. Tick the box that best describes your child's situation. If you find some of the questions difficult to answer, we still ask you to please try to answer them.

*If you respond with "Yes, earlier", please also indicate at what age the respective problem or issue started and at what age it stopped.*

*If you respond with "Yes, now", please also indicate at what age the respective problem or issue started.*

| | *No, never* | *Yes, now* | *Yes, earlier* | *Started (age):* | *Stopped (age):* |
|---|---|---|---|---|---|
| **1.** Do you think your child has or has had problems with eating characterized by avoidance or restriction of foods (i.e., that your child eats only a small range of foods or very little overall)? | ☐ | ☐ | ☐ | | |
| **2.** Has any health professional (at preschool/school, BVC, or other health care) said that your child has problems with avoidant or restrictive eating? | ☐ | ☐ | ☐ | | |

*If you replied "No, never" to* ***both*** *question 1* ***and*** *2, please continue with question 14.*

| | *No, never* | *Yes, now* | *Yes, earlier* | *Started (age):* | *Stopped (age):* |
|---|---|---|---|---|---|
| **3.** Have your child's eating habits led to your child losing weight, not gaining weight or not growing taller as they should? | ☐ | ☐ | ☐ | | |
| **4.** Has any health professional said that your child has nutritional deficiencies due to their eating habits (e.g., vitamin or iron deficiency)? | ☐ | ☐ | ☐ | | |
| **5.** Has your child been prescribed dietary supplements containing vitamins and/or minerals to address nutritional deficiencies? | ☐ | ☐ | ☐ | | |
| **6.** Has your child required high-calorie supplements (e.g., nutritional drinks) to be able to maintain or gain weight? | ☐ | ☐ | ☐ | | |
| **7.** After the age of 6 months, has your child required tube feeding (food or fluid via a tube in the nose or into the stomach) to maintain proper nutritional status? | ☐ | ☐ | ☐ | | |
| **8.** Do your child's eating habits negatively affect their functioning almost daily (e.g., in preschool/school, activities with family/friends)? | ☐ | ☐ | ☐ | | |

*Please answer question 9 only if your child is 6 years or older:*

| | *No, never* | *Yes, now* | *Yes, earlier* | *Started (age):* | *Stopped (age):* |
|---|---|---|---|---|---|
| **9.** Has your child ever restricted their eating because they wanted to lose weight or because they were afraid of gaining weight? | ☐ | ☐ | ☐ | | |

**10.** Do you suspect or know that your child's eating problems are primarily due to a medical condition or a mental disorder? ☐ No ☐ Yes *If yes, which one(s)?* ____________

| | *No, never* | *Yes, now* | *Yes, earlier* | *Started (age):* | *Stopped (age):* |
|---|---|---|---|---|---|
| **11.** Does your child often avoid eating foods with certain smell, taste, appearance, temperature, or consistency/texture (e.g., crispy or soft)? | ☐ | ☐ | ☐ | | |
| **12.** Does your child often avoid eating foods because they are worried about e.g., choking, vomiting/being sick, tummy aches, diarrhea, or an allergic reaction? | ☐ | ☐ | ☐ | | |
| **13.** Does your child often eat too little because of low interest in eating and/or low appetite? | ☐ | ☐ | ☐ | | |

**14.** Has your child ever received a diagnosis because of difficulties with eating? ☐ No ☐ Yes *If yes, which one?* ____________

**15.** What is your relationship to the child? ☐ Mother ☐ Father *If other, who?* ____________

*© Lisa Dinkler, Rachel Bryant-Waugh, Katarzyna Brimo, Maria Råstam, Christopher Gillberg (Sept 2020)*

**Table S1** Item-by-item comparison of the original screener for Avoidant/Restrictive Food Intake Disorder (ARFID) and the refined version, the ARFID Brief Screener (ARFID-BS)

| **Original ARFID screener** | | | **ARFID-BS** | | | |
|---|---|---|---|---|---|---|
| **Criterion** | **Item** | **Response options** | **Criterion** | **Item** | **Response options** | **Explanation of modifications** |
| **Diagnostic criteria** | | | | | | |
| A | 57. Do you think your child has an eating or feeding disturbance characterized by avoidance or restriction of food intake? (avoidance and restriction can relate to the range of foods eaten as well as the overall amount eaten) | No, never<br>Yes, currently<br>Yes, earlier | A-a | 1. Do you think your child has or has had problems with eating characterized by avoidance or restriction of foods (i.e., that your child eats only a small range of foods or very little overall)? | No, never<br>**Yes, now**<br>**Yes, earlier**<br>Started (age):<br>Stopped (age): | Simplified wording to make the question easier to understand for parents |
| | | | A-b | 2. Has any health professional (at preschool/school, BVC, or other health care) said that your child has problems with avoidant or restrictive eating? | | Question added as parents might not be of the same opinion (A-a) as health professionals |
| | | | Skipping rule | *If you replied "No, never" to both question 1 and 2, please continue with question 14.* | | Skipping rule introduced in order to reduce burden of responding to the questionnaire for parents of children without eating problems |
| A1 | 58. Over the past 3 months has there been concern that your child has not gained weight or grown as he/she should? | No, never<br>Yes, currently<br>Yes, earlier | A1 | 3. Have your child's eating habits led to your child losing weight, not gaining weight or not growing taller as they should? | No, never<br>**Yes, now**<br>**Yes, earlier**<br>Started (age):<br>Stopped (age): | Added relation to eating habits as the cause of weight/growth problems; removed limitation "over the past 3 months", otherwise criterion A1 could not be applied to determine previous ARFID/lifetime prevalence |

**Table S1** (continued)

| Original ARFID screener | | | ARFID-BS | | | |
|---|---|---|---|---|---|---|
| **Criterion** | **Item** | **Response options** | **Criterion** | **Item** | **Response options** | **Explanation of modifications** |
| A2 | 59. Has your child been identified by a health professional as having any nutritional deficiency? | No, never<br>Yes, currently<br>Yes, earlier | A2 | 4. Has any health professional said that your child has nutritional deficiencies due to their eating habits (e.g., vitamin or iron deficiency)? | No, never<br>**Yes, now**<br>**Yes, earlier**<br>Started (age):<br>Stopped (age): | Added that nutritional deficiencies are related to eating habits; examples of nutritional deficiency added |
| A3-a | 60. Has your child been prescribed dietary supplements (e.g., vitamins, minerals) to address nutritional deficiencies? | | A3-a | 5. Has your child been prescribed dietary supplements containing vitamins and/or minerals to address nutritional deficiencies? | | *(minimal)* |
| A3-b | 61. Did your child ever need nutritional supplement drinks (or other high-energy drinks) to be able to maintain/gain weight? | | A3-b | 6. Has your child required high-calorie supplements (e.g., nutritional drinks) to be able to maintain or gain weight? | | *(minimal)* |
| A3-c | 25. My child has required supplemental tube feeds to maintain proper nutritional status. *(from BPFAS;* ***removed from the diagnostic algorithm due as response options provide no indication of timing)*** | Never<br>Rarely<br>Sometimes<br>Often<br>Always | A3-c | 7. After the age of 6 months, has your child required tube feeding (food or fluid via a tube in the nose or into the stomach) to maintain proper nutritional status? | | Added time restriction, in order to reduce risk of false negatives e.g. children who required tube feeding only during the newborn period; added explanation of tube feeding |
| A4-a | 63. Do you believe that your child's current eating pattern causes any distress for your child? | Not at all<br>Yes, somewhat<br>Yes, a lot | | | | Item removed: assessment of impairment in functioning according to ICD-11 deemed sufficient with the following A4 question |
| A4-b | 65. Does your child's current eating pattern interfere with his/her social functioning (e.g. attending preschool, affecting meals in preschool, making friends, play, activities)? | | A4 | 8. Do your child's eating habits negatively affect their functioning almost daily (e.g., in preschool/school, activities with family/friends)? | No, never<br>**Yes, now**<br>**Yes, earlier**<br>Started (age):<br>Stopped (age): | Severity rating included into the item ("almost daily") |

**Table S1** (continued)

| Original ARFID screener | | | ARFID-BS | | | |
|---|---|---|---|---|---|---|
| **Criterion** | **Item** | **Response options** | **Criterion** | **Item** | **Response options** | **Explanation of modifications** |
| C | 51. My child says that he/she feels fat, even if other people do not agree with him/her. | Never<br>Rarely<br>Sometimes<br>Often<br>Always | | | | Item removed: interpretation uncertain (children might mimic saying something like this without really meaning it) |
| | | | C | *Please answer question 9 only if your child is 6 years or older:* 9. Has your child ever restricted their eating because they wanted to lose weight or because they were afraid of gaining weight? | **No, never**<br>Yes, now<br>Yes, earlier<br>Started (age):<br>Stopped (age): | Item added to assess food restriction due to body image concerns (exclusion criterion); age restriction added as weight/shape concerns unlikely to be relevant for children under 6 years |
| D | 62. If your child has any problems with weight, growth or nutrition, is this primarily due to a current medical problem? Medical problem: | No<br>Yes | D | 10. Do you suspect or know that your child's eating problems are primarily due to a medical condition or a mental disorder? If yes, which one(s)? | **No**<br>Yes | The question now relates directly to problems with eating due to a medical condition instead of relating to problems with weight/growth/ nutrition; mental disorders added as a potential cause of eating problems |
| **Diagnostic algorithm**<br>*ARFID*:<br>A + (A1 or A2 or A3-a or A3-b or A4-a or A4-b) + C + D<br>*ARFID with physical impairment:*<br>at least one of A1, A2, A3-a or A3-b has to be met<br>*ARFID without physical impairment:*<br>A4-a and/or A4-b are met, but not A1, A2, A3-a or A3-b | | | **Diagnostic algorithms**<br>*ARFID:*<br>(A-a or A-b) + (A1 or A2 or A3-a or A3-b or A3-c or A4) + C + D<br>*ARFID with physical impairment:*<br>at least one of A1, A2, A3-a or A3-b has to be met<br>*ARFID without physical impairment:*<br>A4 is met, but not A1, A2, A3-a or A3-b | | | |

**Table S1** (continued)

<table>
<tr><th colspan="3">Original ARFID screener</th><th colspan="3">ARFID-BS</th><th></th></tr>
<tr><th>Criterion</th><th>Item</th><th>Response options</th><th>Criterion</th><th>Item</th><th>Response options</th><th>Explanation of modifications</th></tr>
<tr><td colspan="7">ARFID Profiles</td></tr>
<tr><td>Lack of interest in eating</td><td>3. My child enjoys eating. (reverse item; from BPFAS)</td><td rowspan="3">Never<br>Rarely<br>Sometimes<br>Often<br>Always</td><td>Lack of interest in eating</td><td>11. Does your child often eat too little because of low interest in eating and/or low appetite?</td><td rowspan="3">No, never<br>Yes, now<br>Yes, earlier<br>Started (age):<br>Stopped (age):</td><td>added that lack of interest in eating actually leads to food avoidance</td></tr>
<tr><td>Sensory sensi-tivity</td><td>54. My child dislikes to eat food with a specific smell, taste, appearance, temperature, or a certain consistency/texture (e.g., crispy or soft).</td><td>Sensory sensitivity</td><td>12. Does your child often avoid eating foods with certain smell, taste, appearance, temperature, or consistency/texture (e.g., crispy or soft)?</td><td>added that sensory sensitivities actually lead to food avoidance</td></tr>
<tr><td>Fear of aversive conse-quences</td><td>53. My child is afraid of eating because of worries about what might happen (e.g., choking, vomiting, stomach aches, diarrhea, or allergic reactions etc.).</td><td>Fear of aversive conse-quences</td><td>13. Does your child often avoid eating foods because they are worried about e.g., choking, vomiting/being sick, tummy aches, diarrhea, or an allergic reaction?</td><td>added that fear of aversive consequences of eating actually leads to food avoidance</td></tr>
<tr><td colspan="7">Feeding or eating disorder diagnosis</td></tr>
<tr><td colspan="4"></td><td>14. Has your child ever received a diagnosis because of difficulties with eating? If yes, which one?</td><td>No<br>Yes</td><td>added in order to be able to validate the questionnaire internally</td></tr>
</table>

BPFAS, Behavioral Pediatric Feeding Assessment Scale

For reasons of simplification, the response options were unified as much as possible in the ARFID-BS. The required response options to meet a criterion are printed in bold. All items are worded as questions (the original screener was a mix of questions and statements). In the ARFID-BS, parents are asked to indicate at which age range each respective problem or issue was present, in order to identify previous ARFID with more certainty than in the original screener, and to gain knowledge on the age of ARFID onset. To screen positive for *current* ARFID, the response needs to be "Yes, now" to at least one of (A-a or A-b) *and* at least one of (A1 or A2 or A3-a or A3-b or A3-c). To evaluate screening status for *previous* ARFID, indications of the age range when certain problems were present need to be evaluated individually.

**Table S2** Principal component analysis of the Child Eating Behavior Questionnaire (CEBQ) with five components

| **Items** | **Components determined through PCA** | | | | | **Explained Variance** | **Cronbach's alpha** |
|---|---|---|---|---|---|---|---|
| | 1 | 2 | 3 | 4 | 5 | | |
| My child… | Satiety Responsiveness | Food Fussiness | Emotional Undereating | Emotional Overeating | Food Responsiveness | | |
| Gets full up easily. | **.410** | | | | | 23.3% | .74 |
| Gets full before his/her meal is finished. | **.408** | | | | | | |
| Leaves food on his/her plate at the end of the meal. | **.366** | | | | | | |
| Has a big appetite. *(reversed)* | **.355** | | | | | | |
| Cannot eat a meal if he/she has had a snack just before. | **.305** | | | | | | |
| Is difficult to please with meals. | .310 | (.120) | | | | 15.1% | .86 |
| Refuses new foods at first. | | **.485** | | | | | |
| Is interested in tasting food he/she hasn't tasted before. *(reversed)* | | **.480** | | | | | |
| Enjoys tasting new foods. *(reversed)* | | **.473** | | | | | |
| Decides that he/she doesn't like a food, even without tasting it. | | **.387** | | | | | |
| Enjoys a wide variety of foods. *(reversed)* | | **.343** | | | | | |
| Eats more when he/she is happy. | | | **.481** | | | 8.0% | .69 |
| Eats less when he/she is tired. | | | **.415** | | | | |
| Eats less when upset. | | | **.396** | | | | |
| Eats less when angry. | | | **.358** | | | | |
| Eats more when annoyed. | | | | **.548** | | 6.5% | .74 |
| Eats more when worried. | | | | **.527** | | | |
| Eats more when anxious. | | | | **.522** | | | |
| Eats more when he/she has nothing else to do. | | | | (.168) | .430 | | |
| Would always have food in his/her mouth if given the chance. | | | | | **.581** | 5.0% | .65 |
| Is always asking for food. | | | | | **.498** | | |
| Would eat too much if allowed to. | | | | | (.227) | | |
| Finds room to eat his/her favorite food even if full up. | | | | | (.208) | | |
| Would eat most of the time if given the choice. | | | | | (.145) | | |

The PCA reproduced the five original CEBQ scales reasonably well, that is, the five components produced by the PCA could be interpreted as the original CEBQ scales *Satiety Responsiveness, Food Fussiness, Emotional Undereating, Emotional Overeating, and Food Responsiveness*. Factor loadings >.30 are displayed. Factor loadings ≤.30 are only displayed (in parentheses) for items that were expected to load >.30 on a specific factor.
